# Moving towards stability: Sleep and cognition across six years of community dance in Parkinson’s disease

**DOI:** 10.64898/2026.08.04.26359697

**Authors:** Simran Rooprai, Ashkan Karimi, Jenna Smith-Turchyn, Nicole D. Anderson, Karolina A. Bearss, Rachel J Bar, David Leventhal, Joseph FX DeSouza

## Abstract

**Background:** Non-motor symptoms, including sleep and cognitive dysfunction, are major contributors to reduced quality of life in people with Parkinson’s disease (PwPD). Dance has been proposed as a promising intervention to improve quality of life in PwPD. Previously, we reported longitudinal trajectories of global cognition following community-based dance; however, little is known about its long-term influence on sleep-related non-motor symptoms and their relationship with global cognitive performance.

**Objective:** We examined the six-year trajectories of sleep and overall non-motor symptom severity among PwPD participating in weekly community-based dance classes compared to a sedentary Reference group. As a secondary objective, we evaluated their association with global cognitive performance as a functional outcome.

**Methods:** This longitudinal observational study followed PwPD engaged in community dance participation as well as a matched sedentary control group from the Parkinson’s Progression Markers Initiative database over six years. Generalized estimating equations (GEE) were used to model group-level trends, with sensitivity analyses conducted to assess the robustness of the findings.

**Results:** Non-motor outcomes showed that insomnia worsened significantly within the Reference group (*p* = .003) but improved among dancers (*p* = .005), with daytime sleepiness remaining stable across both groups. When sleep was used as a predictor of cognition, global cognitive performance trended to improve in the Dance group (*p* = .078) and declined mid-period in the Reference group (*p* = .014). In addition, overall non-motor symptom severity worsened in the Reference group (*p* = .011) but remained stable in the Dance group. Constipation also worsened significantly in the Reference group (*p* = .012) compared to the Dance group.

**Conclusion:** The present study demonstrates that community-based dance may support select non-motor symptoms, including insomnia, and cognitive resilience in PwPD. Findings reinforce dance as a valuable, real-world, non-pharmacological approach to slow functional decline in PD.

## Introduction

Cognitive and non-motor symptoms represent a crucial aspect of Parkinson’s disease (PD). Although traditionally marked by motor signs, such as rigidity, tremors, and bradykinesia, PD is further characterized by several non-motor symptoms and cognitive impairments, which may present years to decades before the emergence of motor symptoms ^1^. Cognitive decline may further evolve into mild cognitive impairment, a significant risk factor for PD-dementia, which affects up to 80% of PwPD over time ^2^. It is estimated that almost all persons with PD (PwPD) will experience some degree of cognitive impairment at some stage of the disease.

Cognitive impairments may include a decline in executive functioning, working memory, attention, visuospatial, or memory domains, and can hinder independence and reduce quality of life, significantly impacting daily functioning ^2^. The inability to understand spatial relationships between objects, known as visuospatial dysfunction, and may lead to a difficulty in carrying out certain activities, including personal hygiene, cooking, feeding, and dressing ^3,4^. Impairments in memory, which are suggested to largely affect recall and recognition ^5,6^, may results in general forgetfulness, slower memory processing, and difficulty with facial recognition among PwPD ^7^. Executive dysfunction, affecting attention and working memory skills, is a hallmark cognitive impairment in PD. PwPD with attentional deficits may struggle to carry out tasks that rely on effortful processing ^8^. For example, Allcock et al. ^9^ noted that an increase in fall risks was associated with attentional deficits in PwPD. Deficits in working memory, defined as the short-term storage and manipulation of information, are another prevalent impairment seen among PwPD ^10^. Given that working memory is a fundamental cognitive skill that intersects with abilities including problem solving and language, a decline in working memory may hinder functional independence ^11^. Executive dysfunction including both attentional and working memory impairments may disrupt the ability to carry out daily tasks, including household chores, finances, and medication management ^12^.

Beyond cognitive impairments, sleep disturbances in PD contribute substantially to disease burden. Insomnia, defined as the inability to fall or remain asleep, and excessive daytime sleepiness (EDS), defined as the inability to stay awake during waking hours, are commonly experienced sleep-related non-motor symptoms, affecting up to 80% and 60% of PwPD, respectively ^13–15^. A longitudinal study by Zhu et al. ^16^ examining excessive daytime sleepiness in PwPD found that 46% of those who did not present with EDS at baseline developed this symptom within 5 years. In another study, Zhu et al. ^17^ showed that approximately 51% of PwPD had developed insomnia at some point across a 5-year period. Furthermore, sleep disturbances are linked to reduced quality of life and are recognized as risk factors for cognitive decline ^18^, while cognitive decline may predict the development of sleep dysfunction as well ^19^. The pathophysiology seen in PD involves the widespread accumulation of Lewy bodies which may impact processes relating to both sleep and cognition, thus leading to sleep and cognitive dysfunction ^20^. Sleep also plays a critical role in cognition ^21^, with sleep dysfunction in healthy individuals linked to cognitive deficits ^20^. Although research on the relationship between cognition and sleep quality in PwPD is limited, previous studies have shown an association between sleep dysfunction and worsening health-related quality of life ^22^, including cognitive impairments ^23^. Goldman et al. ^24^ found that PwPD with worse excessive daytime sleepiness had greater cognitive impairment. With respect to domain specific analyses, Stavitsky et al. ^20^ demonstrated that attentional and executive dysfunction was associated with poor sleep quality in PwPD, whereas the memory domain was not.

In response to the limitations and challenges associated with pharmacological approaches, such as dopaminergic therapies, which mainly address motor impairments ^25^, there is a growing body of literature surrounding innovative non-pharmacological approaches, such as structured community dance programs, to alleviate symptom burden, thus improving quality of life ^26,27^. The complexity of dance, which integrates movement and music along with social engagement, has been recognized as a potential novel approach for slowing down PD progression ^28^. In our previous analysis, we observed improved global cognitive outcomes in PwPD who engaged in weekly dance classes across a six-year period, whereas a sedentary PD-Reference group exhibited no changes ^29^. Building on these findings, the present analysis examines sleep disturbances and overall non-motor symptom burden, as well as their association with changes in global cognition over time following dance. Exploring the relationship between dance participation, global cognition, and select non-motor symptoms may provide greater insights and a richer understanding of community-based interventions to better support PwPD.

The primary objective of this study was to examine longitudinal changes in sleep and overall non-motor symptoms severity between a Dance and sedentary Reference group. Our secondary objective was to examine the impact of overall non-motor symptom severity, including sleep disturbances, on global cognitive performance across time. We hypothesized that volunteers in the Dance group would demonstrate improvements across sleep-related and overall non-motor symptom outcomes, with better sleep and non-motor symptoms associated with improved global cognitive performance, relative to the sedentary Reference group over the six-year study period.

## 2. Methods

The present study utilizes the longitudinal and observational study design and participant cohort previously described in Rooprai et al. ^29^, consisting of 43 volunteers with PD (Dance group; N_male_ = 25; M_age_ = 70.0, SD = 7.0; M_H&Y_ = 1.3, SD = 0.8) enrolled in community-based dance programs in Toronto, Ontario, Canada, as well as 28 sedentary-PwPD (Reference group; N_male_ = 21; M_age_ = 66.7, SD = 7.3; M_H&Y_ = 1.4, SD = 0.8) from the Parkinson’s Progression Markers Initiative (PPMI) database. The PPMI is a longitudinal study funded by the Michael J. Fox Foundation, with the goal of identifying biomarkers for PD. Reference group participants were characterized as sedentary based on their responses on the Physical Activity Scale for the Elderly (PASE), where no engagement in physical activity was reported, including structured (e.g., dance or exercise programs) or non-structured exercises (e.g., walking). The Dance group participated in weekly dance classes based on the Dance for PD method, each lasting 1 hour and 15 minutes, for varying durations across a six-year period (2014-2019; see Bearss et al. ^30^ for the full dance protocol). The study was approved by the Office of Research Ethics (ORE) at York University (approval numbers 2013-211, 2017-296 and 2026-186), and all volunteers in the Dance group provided written informed consent. To avoid redundancy, methodological components that differ from the published protocol are described in detail below.

### 2.1 Outcome Measures

#### Cognition

Volunteers in the Dance group completed the Mini-Mental State Examination (MMSE) ^31^ at one or more time points across the six-year period. Given the observational study design, the number and timing of assessments varied across volunteers; therefore, all available observations were included regardless of the number of assessments completed. For the Reference group, cognitive data was retrieved from the PPMI database for the same study period (2014-2019), where cognitive performance was assessed using the Montreal Cognitive Assessment (MoCA) ^32^. Because the MMSE and MoCA are scored on different scales, scores were standardized by converting raw scores to z-scores to enable between-group analyses (Rooprai et al., ^29^ for the full protocol).

#### Non-Motor and Sleep

For an assessment of non-motor symptoms, aspects of daily living, and PD-related complications, parts I, II, and IV, respectively, of the Movement Disorder Society—Unified Parkinson’s Disorder Rating Scale (MDS-UPDRS) ^33^ were administered at the end of each dance class. The MDS-UPDRS was not administered in 2015 for the Dance group, thus data for that year were unavailable. As with cognitive assessments, the number and timing of the MDS-UPDRS assessments varied across volunteers, therefore, all available observations were included regardless of the number of assessments completed. For the Reference group, MDS-UPDRS data was retrieved from the PPMI database for the same study period (2014-2019). For both groups, sleep-related disturbances were assessed using items 1.7 and 1.8, labelled sleep problems (i.e., insomnia) and daytime sleepiness, respectively, of the MDS-UPDRS Part I Patient Questionnaire, which was self-reported by volunteers. A total sleep score was computed as the sum of sleep problems (i.e., insomnia) and daytime sleepiness to capture overall sleep disturbance.

### 2.2 Statistical Analyses

Standardized global cognitive scores were retrieved from our previously ^29^ used analysis to examine associations with the MDS-UPDRS non-motor outcomes, including sleep disturbances. Higher cognitive scores indicate better cognitive performance. Part 1 of the MDS-UPDRS is an assessment of PD-related non-motor symptoms and is divided into two subsections ^33^. Data for most items in the first subsection were not available for the Dance group, thus only the second subsection (Patient Questionnaire) was analyzed for both groups to maintain comparability. Individual non-motor items from the Patient Questionnaire were examined separately, and a half total score, which represents the sum of all items from this subsection, was used as an indicator of overall non-motor symptom burden. Group-level analyses were also conducted individually for non-motor and sleep-related variables. Higher non-motor and sleep scores indicate worse symptom severity.

Longitudinal changes were modelled using generalized estimating equations (GEE), which account for correlations in repeated measures. GEE was the chosen statistical approach because it provides group-level estimates and accommodates unbalanced data structures in observational study designs ^34^. All primary models included baseline observations as covariates in GEE models to control for between-group imbalances at baseline, with year treated as a categorical variable to accommodate missing and unbalanced observations as well. The year was further collapsed into three periods to improve model stability: Early (2014-2015; *n* = 144 observations, 71 subjects), Mid (2016-2017; *n* = 147 observations, 72 subjects), and Late (2018-2019; *n* = 152 observations, 71 subjects). Models assumed an independence working correlation structure, with specified robust standard errors, as the goal was to obtain population-level estimates, rather than individual-level trajectories. Sensitivity analyses were also conducted to assess the robustness of study findings under alternative assumptions. Linear mixed models (LMM) were employed with year treated as a categorical predictor to analyze trends across time. For cross-domain analyses examining the association of total sleep and overall non-motor symptom severity with cognitive performance, total sleep and Patient Questionnaire scores were included as time-varying covariates in the GEE models. All statistical analyses were performed using JupyterLab (version 3.6.3). All tests were two-tailed with statistical significance set at *p* < 0.05.

## 3. Results

### 3.1 Descriptive Statistics

Table 1 provides the baseline descriptive statistics for demographic and clinical variables. At baseline, the Dance and Reference groups did not differ significantly in disease severity, as measured by the Hoehn & Yahr scale, and UPDRS Part I (Patient Questionnaire) scores, including total sleep. Standardized global cognition differed slightly at baseline, with higher scores observed in the Dance group.

**Table 1.** Descriptive statistics for demographic, clinical, cognitive, and non-motor variables in the Dance and Reference groups. All values are presented as mean (SD), except gender, which is presented as F/M. An asterisk indicates statistical significance. *Note.* Raw baseline scores were MMSE = 27.98 ± 2.42 for the Dance group and MoCA = 25.43 ± 3.05 for the Reference group.

| Variable | Dance Group<br>Volunteers (n = 43) | Reference Group<br>Participants (n = 28) | *p*-value |
| --- | --- | --- | --- |
| Mean age | 70.0 (7.0) | 66.7 (7.3) | 0.064 |
| Gender (F/M) | 18/25 | 7/21 | 0.205 |
| Hoehn & Yahr | 1.3 (0.8) | 1.4 (0.8) | 0.609 |
| Standardized<br>Cognition | 0.82 (0.23) | 0.70 (0.20) | 0.023* |
| UPDRS Part I Patient<br>Questionnaire | 7.24 (3.85) | 6.38 (4.62) | 0.166 |
| Total Sleep Score | 2.25 (1.42) | 2.26 (1.64) | 0.311 |

### 3.2 Outcomes of Insomnia and Daytime Sleepiness

Group-level trends of insomnia and daytime sleepiness are presented in Figure 1. At baseline in 2014, the baseline-adjusted GEE model found no significant differences between the Dance and Reference groups for insomnia (β = 0.29, 95% CI [-0.09, 0.67], *p* = .136) or daytime sleepiness (β = -0.07, 95% CI [-0.35, 0.21], *p* = .607). A significant positive effect of year (β = 0.15, 95% CI [0.05, 0.25], *p* = .003) indicated that insomnia worsened over time in the Reference group. The Dance group, however, demonstrated an improvement in insomnia symptoms over time compared to the Reference group (group x year interaction: β = -0.20, 95% CI [-0.35, - 0.06], *p* = .005). For daytime sleepiness, neither group demonstrated significant change across the study period (Reference β = -0.006, 95% CI [-0.10, 0.09], *p* = .893, interaction β = 0.07, 95% CI [-0.07, 0.20], *p* = .314), suggesting stability across time in both groups. Results were consistent across the primary baseline-adjusted GEE and sensitivity LMM analyses. For insomnia, both models revealed significant worsening over time in the Reference group. In addition, both models found no significant change in daytime sleepiness between the Dance and Reference groups.

**Figure 1.**
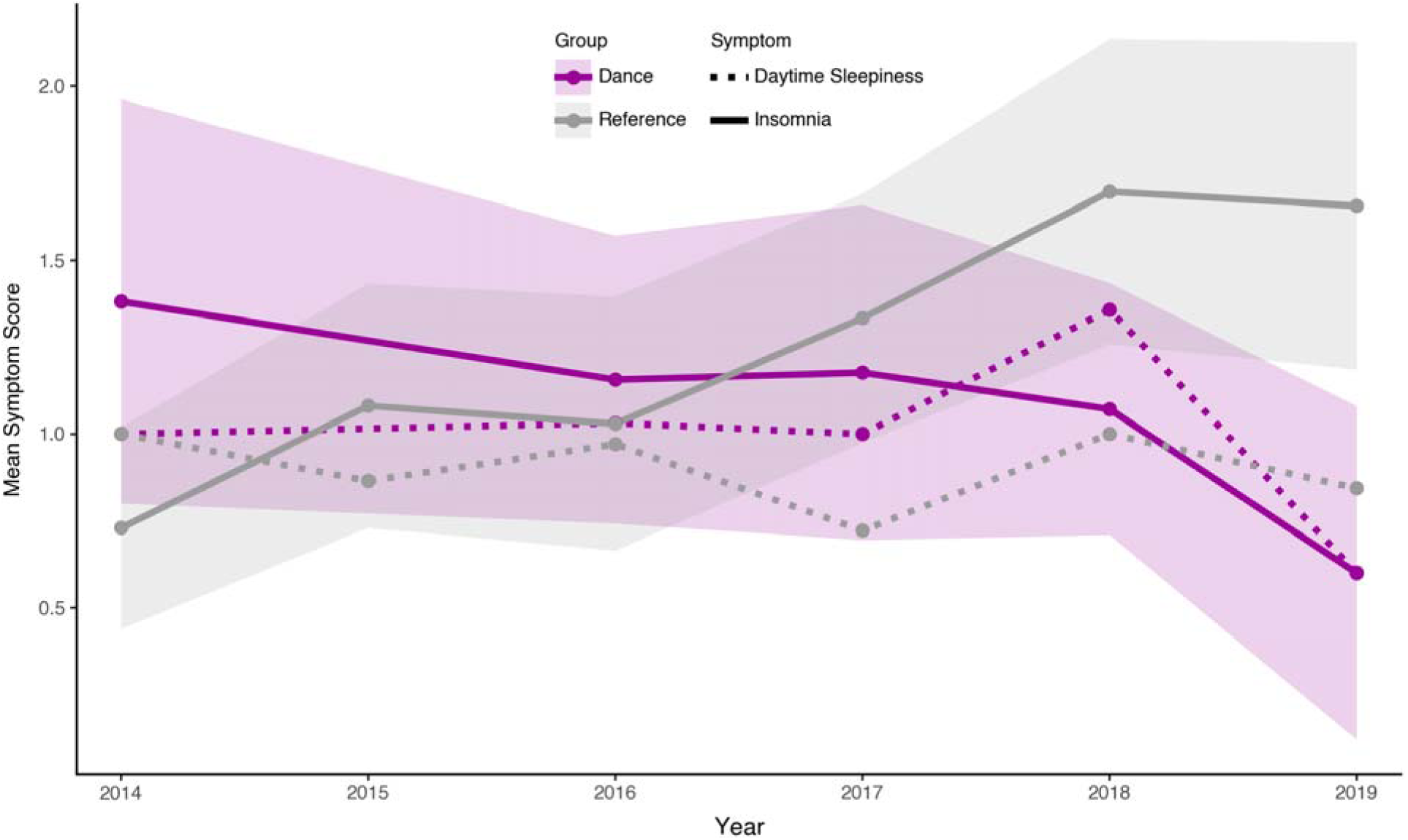
Mean scores of insomnia and daytime sleepiness across six years for the Dance and Reference groups. Line plots display annual group-level trajectories for insomnia and daytime sleepiness from 2014-2019. The Dance group is shown in purple, while the Reference group is shown in grey. Dotted lines are used to represent daytime sleepiness, while insomnia is depicted by solid lines. Higher scores reflect greater symptom severity. Error bars indicate the standard error around the yearly mean score for each group and symptoms. The figure illustrates the significant group x year interaction for insomnia, characterized by improvement in the Dance group and worsening in the Reference group over time.

### 3.3 Overall Non-Motor Symptom Outcomes

Baseline-adjusted GEE models found no significant difference in non-motor symptom burden between the Dance and Reference groups (β = -0.2334, 95% CI [-0.808, 0.341], *p* = .426) at baseline in 2014. The Dance group demonstrated no significant decline in MDS-UPDRS scores across the six-year study period, with no significant change observed during the early to mid (β = -0.26, 95% CI [-1.12, 0.59], *p* = .547), or late periods (β = -0.24, 95% CI [-1.48, 1.01], *p* = .711). A group-by-period interaction revealed a trend towards higher symptom burden in the Reference group during the Mid period (β = 1.06, 95% CI [-0.10, 2.21], *p* = .073) and significant worsening in the late period (β = 2.45, 95% CI [0.55, 4.35], *p* = .011) compared with the Dance group (see Figure 2). A LMM was used to compare the robustness of the GEE findings. No significant group difference was observed at baseline (*p* = .35) and no significant main effects of period were detected (all *p* > .68), aligning with the primary analysis. Instead, a significant interaction effect was observed during the Late period, indicating higher MDS-UPDRS non-motor symptom severity in the Reference group relative to the Dance group (*p* = .005). The interaction at the Mid period showed a similar trend, although this did not reach statistical significance (*p* = .14). Findings of the sensitivity analysis were consistent with the primary GEE results, demonstrating a gradual increase in non-motor symptom burden within the Reference group, while the Dance group remained stable across the six-year study period. Most individual non-motor symptoms remained stable across time within the Dance group and between both the Dance and Reference groups (all *p* > 0.05). However, two significant group-by-period interactions revealed worsening of insomnia and constipation in the Reference group, when compared to the Dance group, during the later years of the study. Full model outputs of all individual non-motor symptoms are provided in Supplemental Table S1.

**Figure 2.**
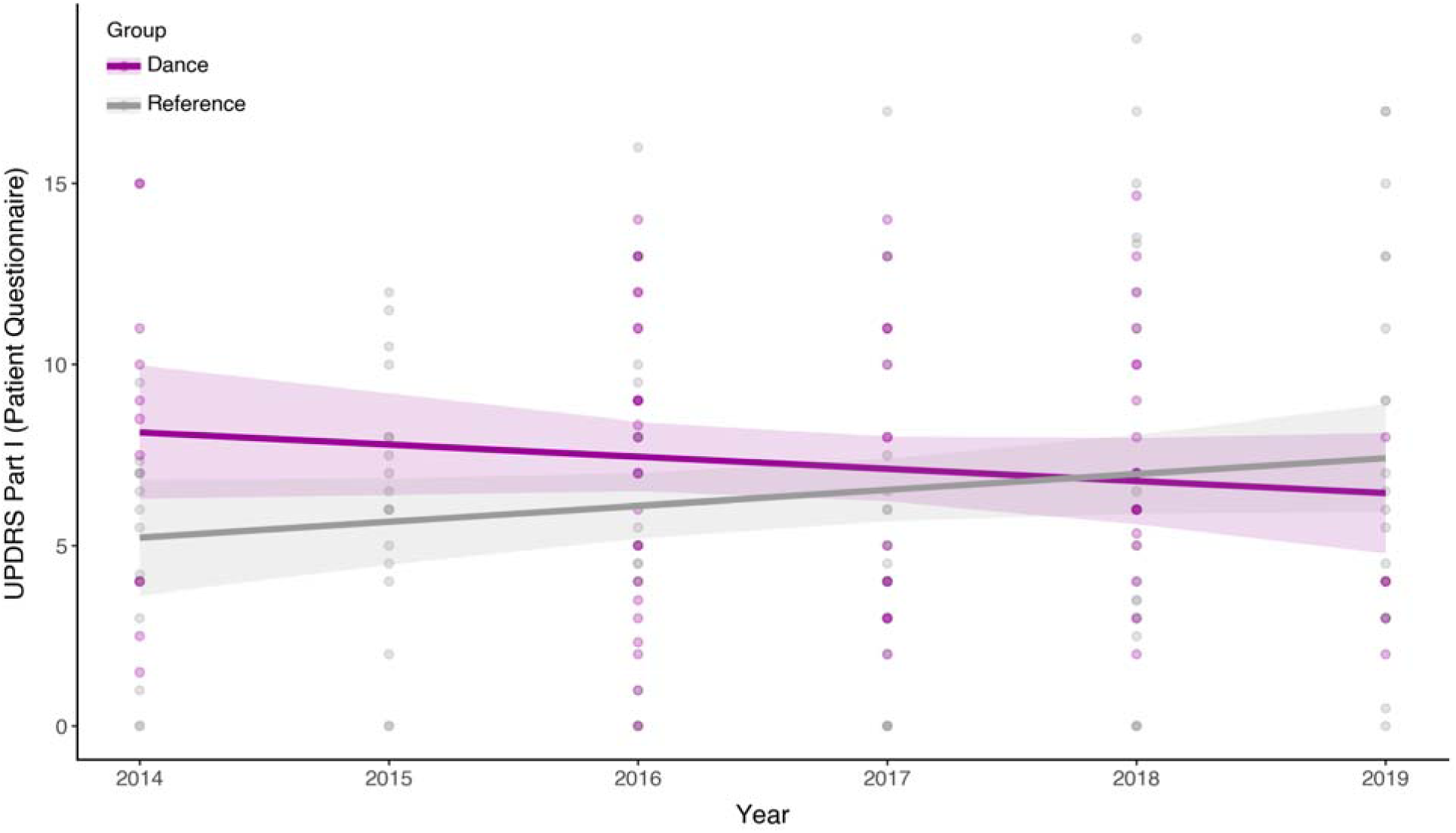
Dance vs Reference Longitudinal UPDRS Part 1 Patient Questionnaire scores from 2014-2019. Scatterplots show individual yearly scores for the Dance and Reference groups, with solid lines depicting fitted linear trends and shaded regions representing the 95% confidence intervals. The Dance group is shown in purple, and the Reference group is shown in grey. Higher scores reflect greater symptom severity. Trends are shown to illustrate overall group-level change across the six-year period.

### 3.4 Cross-Domain Associations

Baseline-adjusted GEE models demonstrated that cognition improved significantly in the Dance group during the Mid period (β = 0.146, 95% CI [0.020, 0.272], *p* = .023) and showed a non-significant upward trend in the Late period (β = 0.0977, 95% CI [-0.011, 0.206], *p* = .078) compared to the Early period. Relative to the Dance group, the Reference group demonstrated a significant decline in cognitive performance during the Mid period (β = -0.1657, 95% CI [-0.298, -0.033], *p* = .014). Although the downward trend continued in the Reference group, it was no longer significant by the Late period (β = -0.104, 95% CI [-0.239, 0.031], *p* = .13). Total sleep was not significantly associated with standardized cognition across groups (β = -0.0033, 95% CI [-0.021, 0.015], *p* = .72), indicating that higher sleep-related symptom severity did not predict poorer global cognitive performance. While worse non-motor symptom severity (MSD-UPDRS Part 1 Patient Questionnaire) was associated with lower global cognitive scores, this relationship did not reach statistical significance (full model output provided in Supplemental Table S2).

Consistent with the baseline-adjusted GEE findings, a LMM found that cognition improved significantly in the Dance group during the Mid and Late periods compared with Early. In comparison, the Reference group demonstrated significantly lower cognitive performance in both the Mid (β = -0.21, 95% CI [-0.34, -0.08], *p* = .001) and Late (β = -0.16, 95% CI [-0.29, -0.02], *p* = .024) periods. Baseline cognition remained a strong positive predictor of subsequent performance (β = 0.68, 95% CI [0.55, 0.81], *p* < 0.001). Total sleep was not associated with standardized cognition (β = 0.001, 95% CI [-0.02, 0.02], *p* = .952).

## 4. Discussion

Across six years of observation, PwPD who engaged in weekly community-based dance classes demonstrated significant improvements and preservation in global cognition relative to baseline, aligning with previous literature demonstrating the cognitive benefits of dance ^35–38^. The high executive demands of dance, particularly stemming from the ability to process, sequence, and recall complex movement sequences timed to music, likely contributed to the cognitive improvements observed in the Dance group ^39,40^ across time and compared to the Reference group. Neuroimaging research further indicates that dance may induce neuroplastic changes, observed among older adults, within the prefrontal cortex, a region that is heavily involved in working memory processes ^41^; thus, suggesting dance may modulate executive functioning across time. Following a 40-week dance intervention, Doi et al. ^42^ also found significant improvements in MMSE scores of older individuals with MCI, aligning with present findings. In contrast, the sedentary Reference group exhibited a mid-period decline in global cognition, like prior evidence suggesting that sedentary behaviour may present as a risk factor for cognitive impairments ^43^. A three-year longitudinal investigation by Jones et al. ^43^, reported that physically active PwPD, or those who became increasingly more active, were at lower risk of severe cognitive decline, whereas those with sedentary lifestyles were at elevated risks for MCI and dementia. Similar findings were demonstrated by Ellingson et al. ^44^ who found that higher engagement in sedentary behaviours were associated with worse self-reported scores on the cognitive subscale of the Parkinson’s Disease Questionnaire-39.

In line with previous literature ^45–47^, insomnia worsened significantly in the Reference group while improving in the Dance group across six-years, leading to a significant group-by-year interaction favouring dance participation. While the pattern of neurodegeneration observed in PD is known to disrupt sleep-related regions in the brain ^48,49^, daytime sleepiness remained stable in both groups across the six-year study period. One explanation may reflect the absence of coexisting sleep disturbances among our groups with mild-PD. For example, a 5-year longitudinal study found that insomnia and daytime sleepiness typically overlap as PD progresses to the advanced stages of the disease ^50^. However, additional research reported a significant association between insomnia and daytime sleepiness among males with mild PD ^51^, suggesting the stability of insomnia in the Dance group, compared with worsening in the Reference group, may reflect the protective role of dance against nocturnal motor disturbances through the preservation of motor symptoms ^26^. Similarly, preliminary findings reported by Krampe et al. ^52^ in a feasibility study on the use of passive bed sensors also found reduced nighttime restlessness in older adults following a 6-week dance intervention compared to a no-dance group. While insomnia was not explicitly analyzed in that study ^52^, brain areas involved in sleep regulation, such as the hypothalamus and limbic regions ^53^ may become engaged through dance. Consistent with the involvement with limbic regions is the association between sustained dance and improved depressive symptoms in PwPD ^54^ Poorer sleep quality was associated with worse depressive symptoms among dancers ^55^, potentially suggesting the link between enhanced emotional regulation and dance in improving sleep quality. Additionally, improvements in insomnia may be explained by the potential interaction between complex exercises, like dance, with the night-time release of melatonin, or reductions in anxiety and depression, mechanisms we aim to explore in future research ^54^.

The existing research on the association between sleep and cognitive dysfunction in PD is inconclusive across the literature ^56,57^. In the present study, we found no relationship between total sleep and cognition in either group, however; Zhong et al. ^58^ similarly reported greater longitudinal decline in global cognition on the MoCA among newly diagnosed and untreated PwPD with insomnia, and no association between MoCA scores and daytime sleepiness. In the present study, the Dance group showed improved or preserved cognitive functioning and stability in insomnia and daytime sleepiness compared to the Reference group, suggesting that the absence of associations may reflect the physical and cognitive benefits of exercise ^59,60^. Consistent with this interpretation, a recent 5-year longitudinal study demonstrated that higher physical activity scores on the PASE were associated with slower deterioration in motor and non-motor symptoms, whereas lower scores predicted faster cognitive and functional decline, including sleep quality, among PwPD ^61^. In a separate analysis of individual non-motor symptom burden, the Reference group demonstrated significant worsening of constipation as well. Interestingly, a recent study by Doi et al. ^62^ investigating the relationship between constipation and nighttime sleep quality among PwPD found that constipation severity was significantly associated with poor sleep in PD. The presence of constipation has been further linked to worse non-motor symptom burden ^63^, with physical inactivity suggested to exacerbate the severity of such non-motor symptoms ^64^, which evidently aligns with our findings demonstrating worse overall non-motor symptom burden in the Reference group. Similarly, the relationship between constipation and disturbed sleep patterns has been previously documented among healthy adults ^65^. One explanation is the negative effects of shorter sleep duration on bowel movements, as shifts in circadian rhythms, which regulate gastrointestinal physiology, may lead to constipation^66^.

### 4.1 Strengths and Limitations

To our understanding, the present study is the first to investigate the influence of community dance engagement on sleep disturbances and overall non-motor symptom severity, alongside their relationship with global cognition, among PwPD across six-years. Specifically, we aimed to analyze insomnia and daytime sleepiness, as well as overall non-motor symptom burden, with global cognition as a functional outcome in a Dance group compared to a sedentary Reference group of PwPD across the same study period. Incorporating a community-based intervention within an observational design spanning six-years provided an opportunity to capture real-world trajectories of cognitive and non-motor symptom progression among PwPD. In contrast to our previous analysis which largely focused on global cognition within the same study sample ^29^, our current findings extend beyond cognition and demonstrate the population-level patterns of non-motor symptomatology in association with long-term dance engagement and physical inactivity, enhancing generalizability to the broader PD community. Although no cognitive change was observed in the Reference group in our previous analysis using the same study sample ^29^, the present baseline-adjusted models may have revealed a time-related decline in cognition, even in the absence of a sleep-cognitive association, potentially strengthening the sensitivity and robustness of our findings.

Despite these strengths, the present study shares several methodological limitations with our previous analysis of global cognition within the same study sample ^29^. Firstly, the study design was observational, and the Dance and Reference groups were non-randomized. Although both groups were matched on key demographic and disease-related variables, unmeasurable confounding variables, such as disease duration, may have played a role. PwPD in the Dance group were not referred or recruited but voluntarily joined the study and were not consistently tested at each time point as well. This variability in the dataset could have reduced statistical power at certain timepoints, preventing subgroup analyses. We did not track volunteer attendance or attrition; therefore, those who discontinued participation may have received reduced benefits compared to individuals who maintained participation, resulting in potential attrition bias. However, a single data point may not reflect the true number of dance classes attended as volunteers may have continued to participate in between the data collection timepoints.

As with our previous analysis ^29^, the lack of consistency between the cognitive tools administered (i.e., MMSE in the Dance group and MoCA in the Reference group) may have introduced differences in measurement sensitivity. Although group-level analyses used standardized cognitive scores, the difference in sensitivity and scoring structures between the two tests could have influenced findings ^67–70^. As such, these methodological limitations should be considered when interpreting the global cognitive outcomes of both groups. While the MMSE is known to exhibit ceiling effects and may lack the ability to detect subtle changes in cognitive functioning ^71,72^, the significant improvement observed in the Dance group suggests that global cognitive gains were sufficiently pronounced to be captured by this assessment tool. Despite the lack of data available on volunteer attendance, the repeated exposure to dance through the longitudinal nature of the intervention may help to support the observed improvements in global cognition. Interestingly, global cognition was assessed using the MoCA in the Reference group where a significant decline was observed, possibly suggesting the improvement in the Dance group may reflect meaningful cognitive change.

Non-motor symptoms, including sleep disturbances, were assessed using the Patient Questionnaire subsection of the UPDRS Part 1. While item 1.7, labelled sleep problems, has been used in previous literature to analyze the severity of insomnia ^50,73^, item 1.8, labelled excessive daytime sleepiness, may not capture the full complexity of this symptom. EDS is a multifaceted symptom that can manifest at various times during waking periods, including while driving or during a conversation ^73,74^, thus a single-item measure may not be suitable for the range of situations in which EDS may be present. As such, future studies should administer comprehensive and descriptive measures, such as the 8-item Epworth Sleepiness Scale, to grasp the functional impact of EDS. Because the Epworth Sleepiness Scale is a subjective and self-reported questionnaire, future studies may also benefit from employing additional measures, including the Multiple Sleep Latency Test and the Maintenance of Wakefulness Test, to objectively monitor and analyze EDS among PwPD ^74,75^. Lastly, the first half of the UPDRS Part I was not collected in the Dance group, limiting our ability to comprehensively assess the full range of non-motor symptoms across both groups.

### 4.2 Conclusion

Taken together, our findings further support dance ^26,27,30,37,47,54^ as a valuable non-pharmacological approach, alongside usual care, to help alleviate non-motor symptom burden ^54^, including sleep and cognitive ^29^ dysfunction, among PwPD. Despite the acknowledged limitations, the present study provides preliminary findings on the longitudinal effects of dance on sleep disturbance, select non-motor symptoms, and their associated with global cognition ^29^. As further research is warranted, our findings may help inform future long-term investigations incorporating more rigorous protocols and comprehensive measures to strengthen alternative models of care, while contributing further to the literature surrounding the benefits of dance for PwPD.

## Acknowledgements

Thanks to all the people from our lab over the years for the project and all the people at our testing locations. We are indebted to S. Leung, R. Cohan, P. Dhami, S. Maguire, D. Rabinovich, H. Tehrani, K McDonald, and R. Andrew for invaluable assistance during all phases of the research program. We are also thankful for the collaborations with Canada’s National Ballet School (A. Seto, A. Powell) and Trinity St Paul’s Church in Toronto, ON, Canada.

PPMI – a public-private partnership – is funded by the Michael J. Safra Foundation, Eli Lilly, Gain Therapeutics, GE HealthCare, Genentech, GSK, Golub Capital, Handl Therapeutics, Insitro, Jazz Pharmaceuticals, Johnson & Johnson Innovative Medicine, Lundbeck, Merck, Meso Scale Discovery, Mission Therapeutics, Neurocrine Biosciences, Neuron23, Neuropore, Pfizer, Piramal, Prevail Therapeutics, Roche, Sanofi, Servier, Sun Pharma Advanced Research Company, Takeda, Teva, UCB, Vanqua Bio, Verily, Voyager Therapeutics, the Weston Family Foundation and Yumanityh, GSK, Golub Capital, Handl Therapeutics, Insitro, Jazz Pharmaceuticals, Johnson & Johnson Innovative Medicine, Lundbeck, Merck, Meso Scale Discovery, Mission Therapeutics, Neurocrine Biosciences, Neuron23, Neuropore, Pfizer, Piramal, Prevail Therapeutics, Roche, Sanofi, Servier, Sun Pharma Advanced Research Company, Takeda, Teva, UCB, Vanqua Bio, Verily, Voyager Therapeutics, the Weston Family Foundation and Yumanity Therapeutics.

## Author Contributions

Simran Rooprai: Formal analysis; Visualization; Writing – original draft. Ashkan Karimi: Visualization. Jenna Smith-Turchyn: Supervision; Writing – review & editing. Nicole D. Anderson: Supervision; Writing – review & editing. Karolina Bearss: Conceptualization; Investigation; Methodology; Project administration. Rachel J Bar: Project administration; Resources. David Leventhal: Resources, Writing – review & editing. Joseph FX DeSouza: Conceptualization; Formal analysis; Funding acquisition; Investigation; Methodology; Project administration; Resources; Supervision; Writing – review & editing.

## Data availability

Data is available on request in accordance with the ethics used at the time of data collection. Data used in the preparation of this article were obtained on [2025–01–05] from the Parkinson’s Progression Markers Initiative (PPMI) database (https://www.ppmi-info.org/access-data-specimens/download-data), RRID:SCR_006431. For up-to-date information on the study, visit http://www.ppmi-info.org/.

## Supplemental Materials

**Table S1.** Baseline-adjusted GEE results for individual non-motor symptoms (UPDRS Part 1 Patient Questionnaire).

| Variable | Predictor | Group(s) | Beta | p-value | 95% CI |
| --- | --- | --- | --- | --- | --- |
| Insomnia | Baseline | Dance vs Reference | -0.0118 | 0.925 | -0.256, 0.232 |
|  | Mid vs Early | Dance | -0.0839 | 0.506 | -0.331, 0.163 |
|  | Late vs Early | Dance | -0.1629 | 0.343 | -0.500, 0.174 |
|  | Group x Mid | Dance vs Reference | 0.2245 | 0.197 | -0.116, 0.566 |
|  | Group x Late | Dance vs Reference | 0.6775 | **0.018** | 0.119, 1.236 |
| Daytime Sleepiness | Baseline | Dance vs Reference | -0.0389 | 0.673 | -0.220, 0.142 |
|  | Mid vs Early | Dance | 0.0741 | 0.402 | -0.099, 0.248 |
|  | Late vs Early | Dance | 0.1836 | 0.308 | -0.170, 0.537 |
|  | Group x Mid | Dance vs Reference | -0.1189 | 0.359 | -0.373, 0.135 |
|  | Group x Late | Dance vs Reference | -0.0811 | 0.743 | -0.565, 0.403 |
| Pain/Other Sensory | Baseline | Dance vs Reference | -0.0746 | 0.617 | -0.367, 0.218 |
|  | Mid vs Early | Dance | -0.2182 | 0.175 | -0.533, 0.097 |
|  | Late vs Early | Dance | -0.2358 | 0.297 | -0.679, 0.207 |
|  | Group x Mid | Dance vs Reference | 0.2411 | 0.237 | -0.158, 0.640 |
|  | Group x Late | Dance vs Reference | 0.2778 | 0.339 | -0.291, 0.847 |
| Urinary Problems | Baseline | Dance vs Reference | 0.1012 | 0.147 | -0.035, 0.238 |
|  | Mid vs Early | Dance | 0.0966 | 0.259 | -0.071, 0.264 |
|  | Late vs Early | Dance | 0.0794 | 0.588 | -0.208, 0.367 |
|  | Group x Mid | Dance vs Reference | -0.109 | 0.454 | -0.395, 0.177 |
|  | Group x Late | Dance vs Reference | 0.1677 | 0.448 | -0.266, 0.601 |
| Constipation | Baseline | Dance vs Reference | -0.0178 | 0.75 | -0.127, 0.092 |
|  | Mid vs Early | Dance | 0.0312 | 0.822 | -0.241, 0.303 |
|  | Late vs Early | Dance | -0.2724 | 0.033 | -0.523, -0.021 |
|  | Group x Mid | Dance vs Reference | 0.0943 | 0.576 | -0.236, 0.425 |
|  | Group x Late | Dance vs Reference | 0.5709 | **0.012** | 0.127, 1.015 |
| Light Headedness | Baseline | Dance vs Reference | -0.011 | 0.935 | -0.276, 0.254 |
|  | Mid vs Early | Dance | -0.075 | 0.455 | -0.272, 0.122 |
|  | Late vs Early | Dance | -0.0791 | 0.568 | -0.351, 0.193 |
|  | Group x Mid | Dance vs Reference | -0.0875 | 0.531 | -0.361, 0.186 |
|  | Group x Late | Dance vs Reference | 0.2188 | 0.348 | -0.239, 0.676 |
| Fatigue | Baseline | Dance vs Reference | 0.024 | 0.85 | -0.224, 0.272 |
|  | Mid vs Early | Dance | -0.1582 | 0.134 | -0.365, 0.049 |
|  | Late vs Early | Dance | 0.036 | 0.779 | -0.216, 0.288 |
|  | Group x Mid | Dance vs Reference | 0.2451 | 0.126 | -0.069, 0.559 |
|  | Group x Late | Dance vs Reference | 0.1342 | 0.5 | -0.256, 0.524 |

The table summarizes model estimates for each symptom, including predictor term, group(s) examined, beta coefficients, p-values, and 95% confidence intervals.

**Table S2.** Standardized Cognition Predicted by UPDRS Part 1 Patient Questionnaire (Between Groups).

| Variable | Predictor | Group(s) | Beta | p-value | 95% CI |
| --- | --- | --- | --- | --- | --- |
| Cognition | Baseline | Dance vs Reference | 0.0281 | 0.516 | -0.057, 0.113 |
|  | Mid vs Early | Dance | 0.1415 | **0.028** | 0.016, 0.267 |
|  | Late vs Early | Dance | 0.0943 | 0.085 | -0.013, 0.202 |
|  | Group x Mid | Dance vs Reference | -0.1614 | **0.016** | -0.293, -0.030 |
|  | Group x Late | Dance vs Reference | -0.0978 | 0.148 | -0.230, 0.035 |
|  | Baseline Cognition | Dance vs Reference | 0.6426 | **<0.001** | 0.484, 0.801 |
|  | UPDRS (centered) | Dance vs Reference | -0.0029 | 0.341 | -0.009, 0.003 |

Displayed values include the predictor term, group(s) examined, beta coefficients, p-values, and corresponding 95% confidence intervals.

